# Higher alpha diversity and Lactobacillus blooms are associated with better engraftment after Fecal Microbiota Transplant in Inflammatory Bowel Disease

**DOI:** 10.1101/2023.01.30.23285033

**Authors:** Yanjia Jason Zhang, Athos Bousvaros, Michael Docktor, Abby Kaplan, Paul A. Rufo, McKenzie Leier, Madison Weatherly, Lori Zimmerman, Le Thanh Tu Nguyen, Brenda Barton, George Russell, Eric J. Alm, Stacy A. Kahn

## Abstract

**Background:** Fecal Microbiota Transplant (FMT) has proven effective in treating recurrent *Clostridioides difficile* infection (rCDI) and has shown some success in treating inflammatory bowel diseases (IBD). There is emerging evidence that host engraftment of donor taxa is a tenet of successful FMT. However, there is little known regarding predictors of engraftment. We undertook a double-blind, randomized, placebo-controlled pilot study to characterize the response to FMT in children and young adults with mild to moderate active Crohn’s disease (CD) and ulcerative colitis (UC).

**Results:** Subjects with CD or UC were randomized to receive antibiotics and weekly FMT or placebo in addition to baseline medications. The treatment arm received seven days of antibiotics followed by FMT enema and then capsules weekly for seven weeks. We enrolled four subjects with CD and 11 with UC, ages 14-29 years. Due to weekly stool sampling, we were able to create a time series of alpha diversity, beta diversity and engraftment as they related to clinical response. Subjects exhibited a wide range of microbial diversity and donor engraftment as FMT progressed. Specifically, engraftment ranged from 26% to 90% at week 2 and 3% to 92% at two months. Consistent with the current literature, increases over time of both alpha diversity (p< 0.05) and donor engraftment (p< 0.05) correlated with improved clinical response. Additionally, our weekly time series enabled an investigation into the clinical and microbial correlates of engraftment at various time points. We discovered that the post-antibiotic but pre-FMT time point, often overlooked in FMT trials, was rich in microbial correlates of eventual engraftment. Greater residual alpha diversity after antibiotic treatment was positively correlated with engraftment and subsequent clinical response. Interestingly, a transient rise in the relative abundance of Lactobacillus was also positively correlated with engraftment, a finding that we recapitulated with our analysis of another FMT trial with publicly available weekly sequencing data.

**Conclusions:** We found that higher residual alpha diversity and Lactobacillus blooms after antibiotic treatment correlated with improved engraftment and clinical response to FMT. Future studies should closely examine the host microbial communities pre-FMT and the impact of antibiotic preconditioning on engraftment and response.

## Background

Fecal microbiota transplant (FMT) is the transfer of healthy fecal microbial communities to a patient with an illness associated with gut microbiome perturbances. The best example to date is the use of FMT to treat *Clostridioides difficile* infection, which has proven to be both safe and highly effective for most patients [1–3]. This success has driven an interest in identifying other diseases where FMT may be beneficial. Perturbations of the microbiome contribute to the pathogenesis of IBD, making it a promising candidate for FMT [4].

Data from several studies, including recent meta-analyses, have demonstrated the efficacy of FMT in some patients with ulcerative colitis (UC) [5–15]. However, the factors that determine responsiveness to FMT are poorly understood. Based on the preliminary data collected, researchers have proposed several theories. Among the proposed factors are the patient characteristics, patient and donor microbial compositions and diversity, degree of donor engraftment, and timing and duration of engraftment [7,12,16,17]. Deeper analyses into the determinants of engraftment have shown highly variable microbe-specific, disease-specific and recipient-specific dynamics. Thus, a generalizable “rule book” for engraftment remains elusive [6,7,12,16,17].

Given that the mechanisms and predictors of success remain unknown, there are no standard pre-conditioning, treatment, or delivery regimens for FMT in IBD. Studies have employed various pre-conditioning regimens including: no bowel prep, bowel prep with laxatives, dietary changes, proton pump inhibitor use, as well as narrow-spectrum and broad-spectrum antibiotics [16,18,19]. There is some suggestion that antibiotic pre-treatment is favorable for both engraftment and clinical response, though the evidence relies on challenging cross-study comparisons [13,18,20]. Furthermore, other studies have reached conflicting conclusions. Our group recently demonstrated that antibiotic pre-conditioning decreased engraftment after FMT, though in a different disease [21]. The literature also provides conflicting answers for even simple questions, such as the effect of recipient alpha diversity on donor engraftment.

In this small study of FMT in adolescents and young adults with IBD, we present a weekly microbiome time series. Frequent sampling allowed us to analyze changes in alpha diversity leading to donor engraftment and thereby identify critical time-points in this FMT protocol. With a specific focus on the post-conditioning, pre-FMT time-point, we show that increased diversity and increased abundance of specific taxa in the family Lactobacilliaceae are correlated with engraftment and clinical response.

## Methods

### Study Design

We conducted a single-center, randomized, double-blind, placebo-controlled trial of FMT in patients with colonic or ileocolonic Crohn’s Disease (CD) and ulcerative colitis (UC). Subjects were recruited from the Boston Children’s Hospital IBD Center as well as through referrals from providers across the country. Standard anthropometric data, past medical and surgical history, and medication history were abstracted from participants ‘medical records.

The primary objective of this study was to assess the safety and tolerability of FMT compared to placebo in pediatric and young adult patients (ages 5-30) with IBD that failed first-line maintenance therapy. The secondary objectives were to identify biomarkers in both donors and recipients that correlate with clinical response.

### Eligibility

Eligible patients were aged 5-30 years with mild to moderate disease activity. Mild to moderate CD Disease Activity was defined as Pediatric Crohn’s Disease Activity Index (PCDAI) >10 but ≤30; mild to moderate UC was defined as Pediatric Ulcerative Colitis Activity Index (PUCAI) > 9 but < 30. Additional eligibility criteria included the presence of visual or histologic evidence of inflammation no more than 105 days before randomization; negative test results for Hepatitis B (HBV), Hepatitis C (HCV), and Human Immunodeficiency Virus (HIV); negative urine pregnancy test for people of childbearing potential; ability to swallow antibiotic, FMT or placebo capsules; and the absence of any known food allergy.

Exclusion criteria included extensive and severe CD (i.e. fistulizing disease, abscess, small bowel obstruction, fevers); patients with recent (within four weeks) dosage changes of biologics, 5-ASA, steroids or immunomodulators; toxic megacolon; known drug allergy to vancomycin, metronidazole or polymyxin; history of aspiration, gastroparesis, surgery involving the upper gastrointestinal tract (that might affect upper gastrointestinal motility) or unable to swallow pills; esophageal dysmotility or swallowing dysfunction; known food allergies; unable or unwilling to receive a retention enema for purposes of induction therapy; recent (within six weeks) systemic antibiotic use; testing consistent with active clostridium difficile; and known prior experience with FMT.

Subjects were maintained on their standard of care medications at the primary provider’s discretion. Patients with mild to moderate disease activity were consented and randomized to receive treatment with antibiotics followed by FMT or placebo. This study was approved by the Boston Children’s Hospital Institutional Review Board and was registered on https://clinicaltrials.gov (Identifier: NCT02330653).

### Randomization

Subjects enrolled were randomized in a 1:1 ratio according to a pre-determined block randomization procedure to receive either the treatment arm or the placebo arm. An unblinded member of the study team maintained the randomization list, ensured subjects were appropriately randomized, and dispensed the correct treatment.

### Study Groups

Subjects in the treatment arm (hereafter referred to as the FMT arm) received seven days of antibiotic pre-treatment beginning on Day -8. One capsule containing metronidazole (weight-dependent dosing, maximum dose of 500 mg) was administered twice a day. Capsules containing 125 mg of vancomycin and 62.5 mg of polymyxin were administered three times a day. The number of capsules given was based on Body Surface Area parameters.

Approximately 48 hours after the discontinuation of antibiotic pre-treatment, on Day 0, subjects were given an induction retention enema of FMT over the course of 15-30 minutes. They were encouraged to retain the fecal matter for as long as possible. Subjects were then observed for at least 60 minutes before discharge. Subjects were subsequently treated with a weekly dose of 30 FMT capsules for the next seven weeks.

FMT material was obtained from OpenBiome in Cambridge, MA using established protocols [12,22]. OpenBiome is a non-profit stool bank dedicated to treating and researching the microbiome. Their work focuses on providing safe and affordable FMT material, thereby removing logistical barriers for patients and physicians.

Subjects in the placebo arm received seven days of corresponding placebo capsules in place of antibiotic treatment. At Day 0, these subjects received a placebo retention enema. Subjects were subsequently treated with a weekly dose of 30 placebo capsules for the next seven weeks.

Subjects provided weekly stool samples and PCDAI/PUCAI surveys to monitor disease activity regardless of the assigned arm. At Week 4 and Week 8, routine laboratory assessments were collected to measure clinical response.

### Open-Label Eligibility and Treatment

After eight weeks of treatment and unblinding, subjects in the FMT arm that responded to treatment and all subjects in the placebo arm were eligible to receive an additional eight weeks of open-label FMT. Response to treatment was defined as a decrease in disease activity index score of at least 10 points or a score of 10 points or less.

### Endpoints/Outcomes

Primary outcome measures included any FMT-related adverse events, grade 2 or above, and the proportion of subjects reporting any FMT-related adverse events of grade 2 or above at eight weeks post-FMT. Patient-related outcomes of abdominal pain and the average daily number of bowel movements were recorded.

Secondary outcome measures included remission as defined by a disease activity index of <10, improvement of inflammatory biomarkers such as fecal calprotectin, erythrocyte sedimentation rate (ESR), C-reactive protein (CRP), changes in gut microbial composition, improvement of disease activity scores (PCDAI ≥ 12.5, PUCAI ≥ 20), and assessment of engraftment of donor microbes into recipients.

### Fecal Sample Collection

Subjects submitted stool samples during screening, baseline, after antibiotics but before FMT, then weekly during blinded and open-label treatment, and during follow-up. These samples were stored at -80°C.

### Microbiome Analysis

We extracted DNA using a Powersoil DNA extraction kit (Qiagen). 16S rDNA libraries were prepared and sequenced by the Broad Institute Genomic Platform, using paired-end 250-bp reads on an Illumina HiSeq. We analyzed 16S data using Qiime2, DADA2, Phyloseq in R, and custom Python scripts [23–25]. We assigned taxonomic labels to 16S sequences using the SILVA database [26]. We used SourceTracker2 to estimate the sources of various bacteria after FMT [27]. We used the pre-antibiotic sample, the post-antibiotics sample, and the known donor as sources for each participant. We also included the other donor, not used in that participant’s FMT, as a negative control. To differentiate the participants who achieved high engraftment from those who did not, we used the q2-sample-classifier in Qiime2, using the Random Forrest classifier [28].

## Results

### Study Enrollment/Patient Characteristics

We selected patients with primarily mild to moderate colonic CD or UC without stricturing or penetrating disease. Four subjects with CD and 11 subjects with UC were consented and randomized to a treatment arm (Figure 1A). The median age at enrollment for CD patients was 20 years (range 18-23) and for UC patients was 24 years (range 14-29). All patients were white, and none were Hispanic or Latino. At baseline, all patients had mild or moderate disease activity index scores. All 4 CD subjects had mildly active disease (PCDAI of 11-25). With respect to subjects with UC, three had mildly active disease (PUCAI scores of 10-24), three had mild to moderate disease (PUCAI scores of 25-39) and three had moderate disease activity (PUCAI scores of 40-69) (Supplemental Table 1).

**Figure 1.**
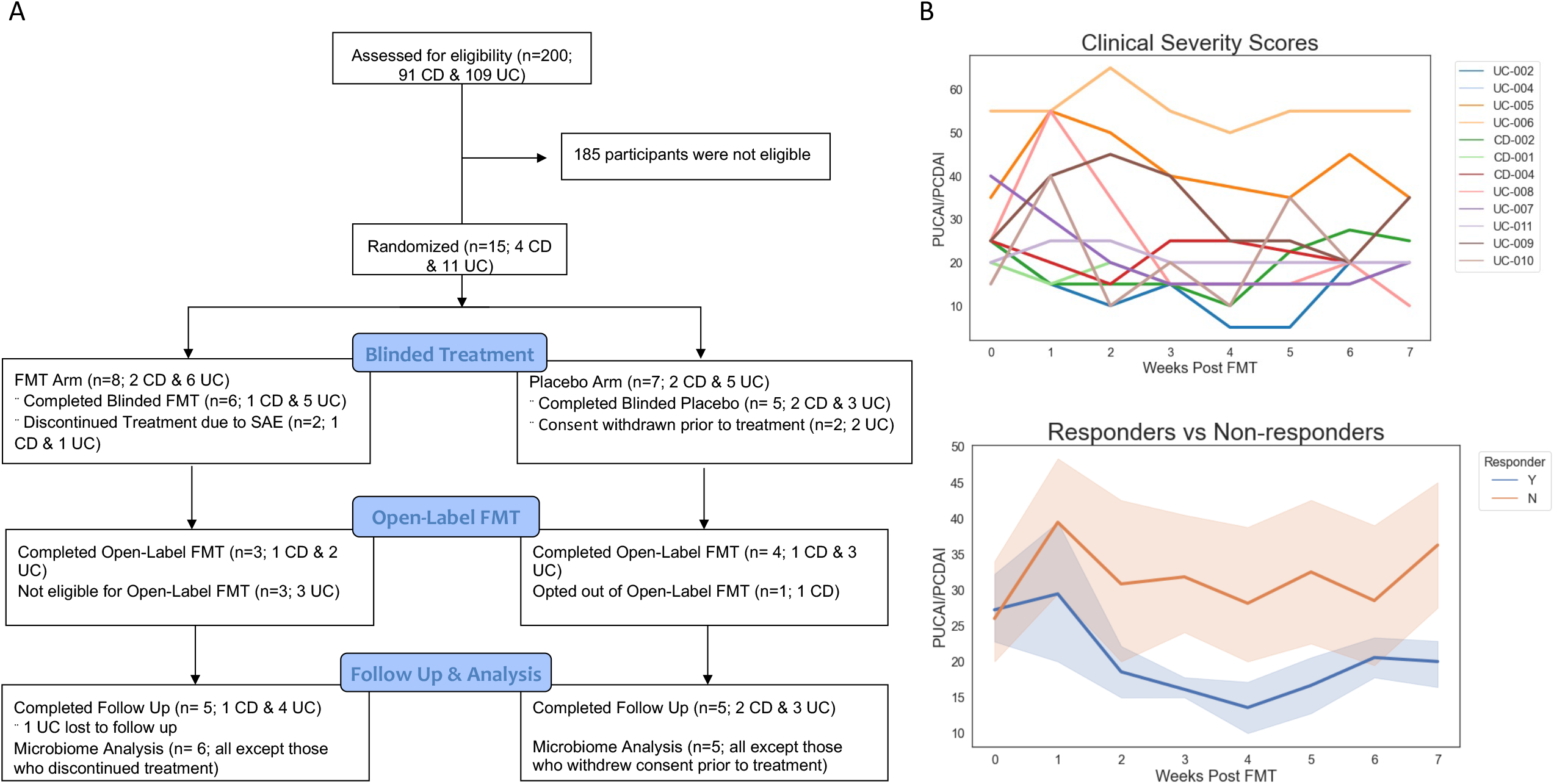
A. Consort Diagram. B. Clinical symptom scores (PUCAI for UC and PCDAI for CD) for individual participants (top) and for responders vs non-responders (bottom).

Eleven (3 CD and 8 UC) subjects completed the blinded phase of the study (Figure 1A). Four (1 CD and 3 UC) subjects were ineligible to continue into the open-label phase because they did not meet response criteria (Figure 1A). Seven (two CD and five UC) subjects completed the open-label phase of the study and six (two CD and four UC) subjects completed long term follow up (Figure 1A).

### Clinical Response

Overall, three patients reported a decrease in IBD-related symptoms. Subjects with UC experienced an average decline in PCDAI score of 5 and an average decline in PUCAI of 7.5 while receiving the experimental arm or during open-label therapy (Figure 1B). The FMT was safe and well-tolerated in a majority of the patients. There were two serious adverse events (SAE): one episode of Grade 3 colitis that was determined to be possibly related to the intervention antibiotic treatment (and not FMT) and one hypersensitivity reaction directly after the FMT induction enema that was deemed probably related. Both subjects were removed from the study. There were other adverse events that were determined to be not related to the study intervention (Supplemental Text).

### Host Microbiome Response

On average, alpha diversity decreased after a week of antibiotics therapy and increased throughout the course of FMT. There were large inter-individual differences in the alpha diversity post-FMT, with Shannon indices ranging from less than 1 to greater than 5 (Figure 2A). We generated a weekly time series of gut microbial changes using serial stool samples for sequencing. The microbial differences between responders and non-responders diverged early.

**Figure 2.**
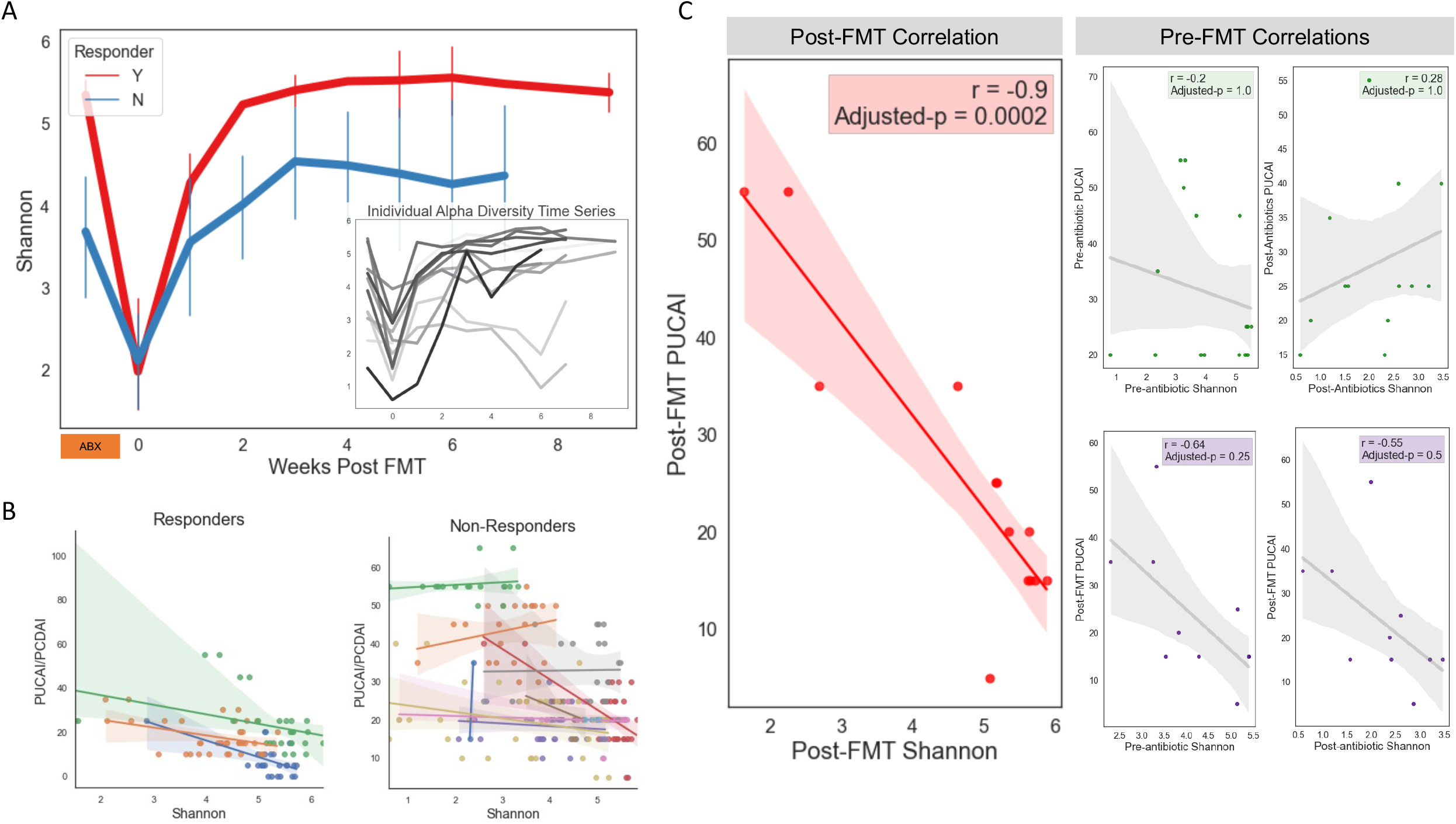
A. Stool alpha diversity (Shannon index) time series for all patients, clinical non-responders (blue) vs. clinical responders (red) and for individual patients, each in a unique shade of gray (inset). B. Correlation between clinical symptom score (PUCAI for Ulcerative Colitis, PCDAI for Crohn Disease) and alpha diversity (Shannon index) for clinical responders (top) and non-responders (bottom). C. Correlation between PUCAI and alpha diversity at the post FMT timepoint (left, red, large panel). Smaller panels: top panels show correlation between PUCAI and alpha diversity at the pre- and post-antibiotic timepoints, bottom panels show correlation between the late post-FMT PUCAI and earlier (pre- and post-antibiotic) alpha diversity metrics.

Starting at two weeks post-FMT, we observed higher alpha diversity in the clinical responders compared to the non-responders (t-test, p-value < 0.05) (Figure 2A). The measured difference in alpha diversity between responders and non-responders was significant even when we used a broader definition of clinical response (Supplemental Figure 1).

We next explored the relationship between alpha diversity and clinical symptoms within individuals. In the three clinical responders we were able to measure a statistically significant relationship (Pearson’s adjusted p-values all < 0.05) between the Shannon Index and PUCAI or PCDAI, where higher diversity correlated with lower disease activity (Figure 2B). We did not observe the same relationship in the non-responders (Figure 2B). Finally, we sought to determine at what point in the treatment course microbial diversity was most correlated with symptom severity. There was no correlation between alpha diversity and disease activity index pre-or post-antibiotic treatment, despite a large range of alpha diversity in the participants (Figure 2C, small panels, top). There was a trend towards higher alpha diversity at earlier time points correlated with lower symptom scores after FMT, but this was not statistically significant (Figure 2C, small panels, bottom). On the other hand, starting at 6 weeks post-FMT, the alpha diversity was significantly correlated with PUCAI or PCDAI (Pearson’s correlation, r = -0.9, adjusted p-value = 0.0002). Higher alpha diversity similarly demonstrated a strong association with lower disease activity (Figure 2C, left panel).

### Donor Engraftment

We hypothesized that the relationship between alpha diversity and clinical improvement was driven largely by donor engraftment. There were 2 FMT donors. Each recipient was randomly assigned to receive capsules from a single donor. The donors were healthy, and their microbiomes were easily distinguishable through 16S sequencing and beta-diversity measurements (Supplemental Figure 2). We measured donor engraftment using SourceTracker2, a Bayesian algorithm based on Gibbs sampling that ultimately assigned a predicted proportion of a stool microbial community that originated from a set of input sources [27]. For each stool sample post-FMT, we used 16S sequences from the recipient’s pre-antibiotic stool sample, the recipient’s post-antibiotic stool sample and the assigned donor’s capsules as potential sources. Additionally, we added the other donor as a dummy source as a way of surveying for source misassignment. On average, the dummy donor was assigned a proportion of 0.016, and there were only 3 of 111 instances where the assigned portion was above 0.05 (Supplemental Figure 3 and Supplemental Table 2).

**Figure 3.**
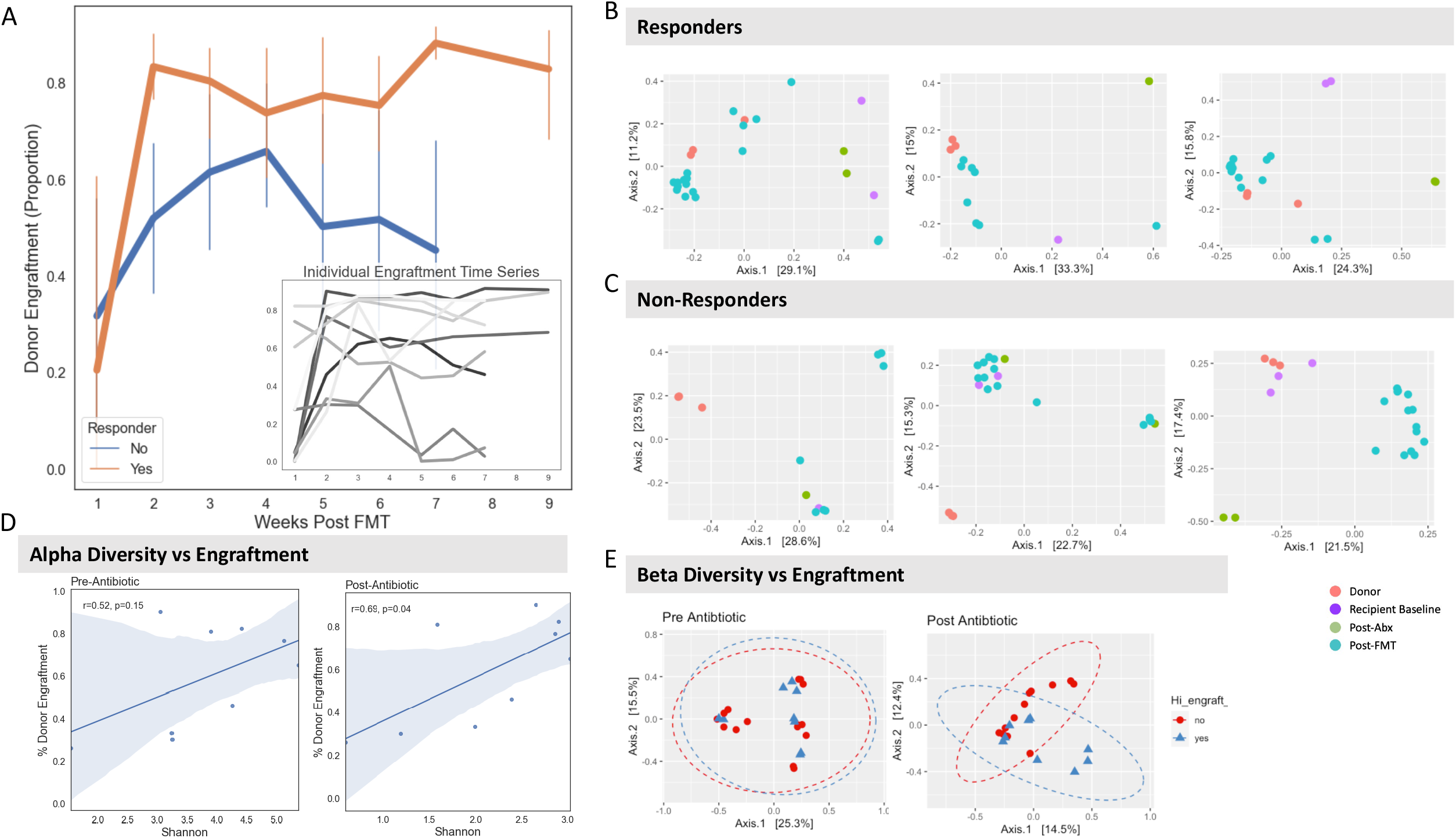
A. Percent Donor Engraftment, as estimated by SourceTracker, in responders vs. non-responders as well as for each individual patient, each in a unique shade of gray (inset) B. Beta diversity plots (Bray-Curtis) for three example responders. C. Beta diversity plots for three example non-responders. D. Correlation between alpha diversity (Shannon Index) and engraftment at the pre-antibiotic (Left) and post-antibiotic (Right) timepoints. E. Beta-diversity plots comparing eventual high engrafters (blue) and low engrafters (red) at the pre-antibiotic (Left) and post-antibiotic (Right) timepoint.

Starting at two weeks post-FMT, responders had higher levels of donor engraftment compared to non-responders (Figure 3A and Supplemental Table 2). This association was statistically significant at week two and week seven post-FMT (T-test, p-value < 0.05). At weeks six through eight, responder microbiome compositions were much closer to their assigned donor than non-responders (Bray-Curtis dissimilarity) (Figure 3B and 3C). In one case, the pre-FMT microbiome was similar to the donor microbiome in a non-responder. However, the participant’s microbiome shifted away from the donor’s microbial profile following FMT. (Figure 3C, right-most panel). In this instance, perhaps antibiotics played as big a role in the resultant microbiome community as the FMT itself.

### Correlates of Engraftment

The degree of engraftment correlated with both restoration of gut microbial diversity and clinical response to FMT. At 3 to 8 weeks post-FMT, there was a wide range of engraftment amongst participants, from less than 0.3% to 91.8% (Figure 3A, inset). We searched for clinical, laboratory and microbial correlates of engraftment by comparing the engraftment at weeks 2 and 7 with other features of the participants. These features included symptom severity scores, inflammatory markers in the stool and blood, as well as the baseline diversity of the gut microbiome before and after antibiotics. The strongest correlate of engraftment at seven weeks post-FMT was engraftment at two weeks post-FMT (Pearson R = 0.69, p-value < 0.05), suggesting that early engraftment—or lack thereof—is predictive of late engraftment (Supplemental Figure 4). The only clinical or laboratory measurement significantly correlated with engraftment was fecal calprotectin. After antibiotics, there was a negative correlation between calprotectin and engraftment (Pearson R = 0.72, p-value < 0.05), suggesting that high levels of gut inflammation may prevent engraftment (Supplemental Figure 4).

Antibiotic pre-conditioning significantly decreases the alpha diversity and changes the ecology of the microbiome (Supplemental Figure 5A and 5B). The unanswered question, however, is whether lower diversity (a “cleaner slate”) or higher residual diversity after antibiotics is a more supportive environment for engraftment. In our small study, we found that higher alpha diversity after antibiotics correlated with higher engraftment (Pearson R = 0.69, p-value < 0.05) (Figure 4D). We found no such correlation between the alpha diversity prior to antibiotics—the participant’s baseline, in other words—and eventual engraftment. Similarly, we were able to separate the post-antibiotic microbial communities in high-engrafting individuals from those in low-engrafting individuals (Bray-Curtis, PERMANOVA p-value < 0.01), whereas pre-antibiotic communities in high engrafting versus low engrafting individuals did not cluster separately (Figure 4E.)

**Figure 4.**
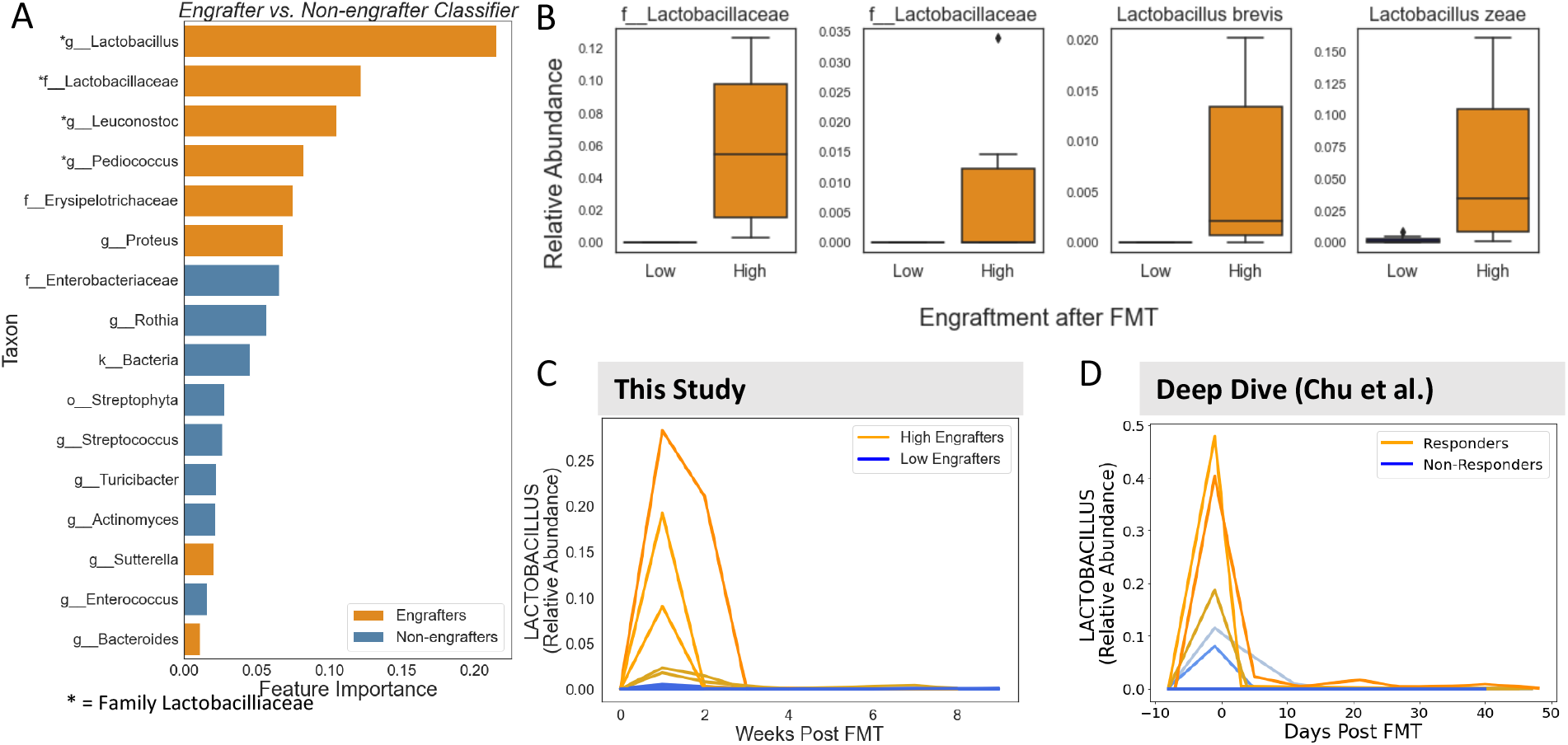
A. Important features (on the genus level) from a random forest classifier using the post-antibiotic microbiome to classify high vs. low engrafters (AUC = 1.00). All taxa are genera, lowest identified taxonomic level is labeled. *Asterisks denote members of the Family Lactobacilliaceae. B. Boxplot of relative abundance (proportion of total reads) of species in the Family Lactobacillaceae that were detected in at least 3 individuals. ***, FDR p-value < 0.001; **, FDR p-value < 0.01; *, FDR p-value < 0.05. C. Time series of relative abundance of genus Lactobacillus (top left) and 3 species within genus Lactobacillus, comparing high (each participant represented in a unique shade of blue) vs. low (each participant represented in a unique shade of orange) engrafters in this study. D. Time series of relative abundance of genus Lactobacillus (top) and 2 species within genus Lactobacillus, comparing responders (each participant represented in a unique shade of blue) vs non-responders (each participant represented in a unique shade of orange) in an independent (n = 6) FMT study in IBD (Chu et al.).

### Members of Family Lactobacilliaceae are Associated with High Donor Engraftment

We next sought to determine if the presence of specific taxa after antibiotic treatment was correlated with engraftment. To that end, we trained a Random Forest classifier that accurately predicted high engraftment in the test set using relative abundance at the genus level. Interestingly, when ranking feature importance, the top 4 features were genera in the family Lactobacilliaceae (Figure 5A). At the timepoint immediately following antibiotics, 4 of the 12 species identified within family Lactobacilliaceae were higher in relative abundance in participants who eventually exhibited high engraftment, including 2 unclassified Lactobacilliaceae species as well as *Lactobacillus zeae* and *Lactobacillus brevis* (Figure 5B). We generated individual taxa time series and compared patients with eventual high versus low engraftment. For the genus Lactobacillus, as well as the species *Lactobacillus zeae, Lactobacillus brevis* and an unclassified Lactobacillus species, we observed a spike in the relative abundance of the taxa immediately following antibiotic therapy (Figure 5C and Supplemental Figure 6). As the FMT (and engraftment) progressed, the Lactobacillus taxa decreased in relative abundance. We are aware of only one other study of FMT in patients with IBD with publicly available 16S sequencing data from the post-conditioning pre-FMT time point [7]. In that study, six patients received FMT, three responded clinically and three did not. Interestingly, we found that the responders had a very similar transient spike in the relative abundance of the Genus Lactobacillus and the two Lactobacillus species that were detected in all six patients in the study by Chu et al. (Figure 5D and Supplemental Figure 6).

## Discussion

The clinical response to FMT in IBD is typically between 20 and 40%, and our study showed similar efficacy [5,8,9,11,13,14]. Three of the 12 patients who completed FMT had a clinical response. In this study, we focused our analysis on what separated eventual success from eventual treatment failure. We found that 1.) engraftment of the donor microbiota was higher in the responders vs. the non-responders, 2.) higher residual alpha diversity after antibiotic therapy was associated with better engraftment and 3.) a relative rise in the abundance of several Lactobacilliaceae taxa after antibiotic therapy correlated with engraftment.

Previous studies have hinted that antibiotic therapy as a component of pre-FMT conditioning improves efficacy, but conditioning regimens remain highly variable, and their effect on engraftment and clinical response is poorly defined [18]. The small size of our study is a significant weakness. However, weekly sampling throughout the course of FMT, as well as specifically sampling the gut microbial community prior to FMT but after antibiotic conditioning, was a strength. The frequent sampling allowed us to study the determinants and kinetics of engraftment. Among the many unanswered questions about the rules of engraftment, we focused on two questions related to kinetics. First, how quickly does a donor microbiome take hold in patients who eventually engraft? Second, at what point prior to FMT are recipient microbial features correlated with eventual engraftment?

In patients who engraft and clinically respond, the engraftment takes root within two weeks. Except for one individual, every participant that achieved engraftment of roughly 75% or greater at the end of the FMT period had greater than 75% engraftment at week 2. The other individual achieved greater than 75% engraftment at week 3. This suggests that features supporting engraftment exist early, perhaps even before the initiation of FMT.

We found no baseline clinical features in our participant pool associated with engraftment. After antibiotics, though, fecal calprotectin was negatively correlated with engraftment, suggesting that decreased gut inflammation after antibiotics supports engraftment. While we could not find any microbial correlates of engraftment at baseline (prior to antibiotics), there were multiple microbial factors *after* antibiotic treatment that correlated with engraftment. We know of only two other studies that sequenced the microbiome after antibiotics but before FMT in patients with IBD. Given the relative importance of this time-limited community to engraftment, we propose that future FMT studies should sample at this point to better understand how to best to prepare a niche for donor engraftment [5,7]. Pre-FMT antibiotics theoretically help eliminate pathogens and commensals that may outcompete donor microbes as they establish their niche in the recipient’s colon. Somewhat contradictory to that theory, our data suggest that maintaining a higher level of diversity after antibiotics predicts better engraftment. While this has not been shown in IBD FMT trials to our knowledge, this is in keeping with two other studies that assessed the determinants of engraftment. In one study of FMT in patients with *Clostridioides difficile* infection, higher recipient alpha diversity was associated with improved donor microbiota engraftment [12]. In a separate study of FMT for patients with irritable bowel syndrome (IBS), antibiotic treatment and the resulting drop in diversity seemed to decrease engraftment after FMT [21]. The discrepant effect of antibiotics in IBD versus IBS (supportive of engraftment in IBD but detrimental in IBS) demonstrates that different diseases may require different pre-FMT conditioning regimens. Perhaps higher pathogen abundance in IBD makes antibiotics more favorable. However, our data shows that there is also high inter-individual variability in the effect of antibiotics, and that even within the same disease, the antibiotics created a supportive milieu for engraftment in some but not in others. Determining who would benefit from antibiotics (and which antibiotics to use) is an area of much-needed continued research.

The final facet of our analysis aimed to determine the specific taxa associated with engraftment. Separating the participants into high versus low engraftment, we looked back at the baseline gut microbiota and the post-antibiotic microbiota for beta diversity analysis. Similar to alpha diversity, we found no measurable differences in the baseline samples. However, there was a clear separation of the high and low engraftment patients at the time point immediately following antibiotics (but before FMT). The main differences were the relative abundance of commensals from the Lactobacilliaceae family. Patients who eventually had high engraftment tended to have higher relative levels of these taxa. Interestingly, in an FMT trial for *Clostridioides difficile* infection, Lactobacilliaceae had the highest dependency score amongst the taxonomic determinants of engraftment identified [12]. Altogether, these findings suggest that Lactobacilliaceae may promote engraftment, perhaps by creating a more favorable niche for the donor microbiome. Multiple species in the genus Lactobacillus have been shown in pre-clinical models to support intestinal regeneration. More specifically, emerging evidence has demonstrated that Lactobacilli can increase goblet cells and mucin production. This could support the engraftment of a new microbiome [29]. At a broader level, these findings suggest that creating an ideal environment for engraftment is key. Improving FMT engraftment depends on our ability to understand—and maybe one day engineer—the microbial and ecological environment into which FMT microbes enter.

## Conclusions

Our study demonstrates the need for more inquiry into recipient characteristics that predict FMT engraftment. By analyzing the large inter-individual differences in engraftment, we propose that higher residual microbial diversity after antibiotics supports engraftment, and that several Lactobacilliaceae taxa may mediate that effect. Ultimately, our study is small and underpowered to fully delineate the determinants of engraftment. We argue that the effects of antibiotic pre-conditioning are still largely unknown, and that methodical characterization (microbial sequencing, deeper clinical phenotyping, multi-omics techniques) of the post-antibiotic (but pre-FMT) state will help determine the factors that make FMT in IBD succeed in some but fail in others.

## Data Availability

All data produced in the present study are contained in the manuscript, available in the supplements, or are available upon reasonable request to the authors.

## List of Abbreviations

CD: Crohn disease;
FMT: Fecal Microbiota Transplant;
PUCAI: Pediatric Ulcerative Colitis Activity Index;
PCDAI: Pediatric Crohn Disease Activity Index;
IBD: Inflammatory Bowel Disease;
IBS: Irritable Bowel Syndrome;
rCDI: recurrent *Clostridiodes difficile* Infection;
UC: Ulcerative Colitis

## Declarations

### Ethics approval and consent to participate

The study was conducted in accordance with the protocol, applicable ICH Guidelines, Good Clinical Practice and the World Medical Association (WMA) Declaration of Helsinki and its amendments concerning medical research in humans.

In accordance with guidelines and U.S. Code of Federal Regulations applicable to clinical studies, the protocol and informed consent/assent forms were reviewed and approved by the Boston Children’s Hospital Institutional Review Board (IRB). The investigator informed the IRB and FDA of subsequent protocol amendments and reportable events as defined by IRB policy and FDA regulation.

All patients and/or their guardians consented to participate in this study.

### Consent for publication

Not applicable

### Availability of data and material

The availability of datasets generated during and/or analyzed during the current study is pending.

### Competing Interests

AB receives research support as a subinvestigator on protocols for Janssen, Abbvie, Takeda, Buhlmann, Arena, Eli Lilly, Bristol Myers Squibb, PROCISE diagnostics. AB receives consulting revenue from Takeda, Best Doctors, Eli Lilly, and Fresenius Kabi. AB received an honorarium from Boston University and Royalties from Up To Date.

No other authors had any competing interests.

### Funding

This research was supported by the Eunice Kennedy Shriver National Institute of Child Health and Human Development (award 5K12HD052896 to Gary R. Fleisher, MD). We would like to thank the Hamel and Rasmussen families for the support of this research.

### Authors’ contributions

YZ performed the microbiome analysis and wrote the manuscript. AB contributed to trial design, performed clinical trial procedures, and critically reviewed the manuscript. EA and TN helped perform the microbiome analysis and edited the manuscript. AK performed clinical data analysis and helped write the manuscript. MD, MW, PR, ML, LZ, BB and GR recruited patients, performed the clinical tasks related to the trial and edited the manuscript. BB was the research manager and supported the clinical trial procedures. GR designed the trial and edited the manuscript. SK was the primary investigator, contributed to clinical trial design, oversaw clinical trial procedures, analyzed clinical data, and was a major contributor in writing the manuscript.

## Figure Legends

**Supplemental Figure 1.**
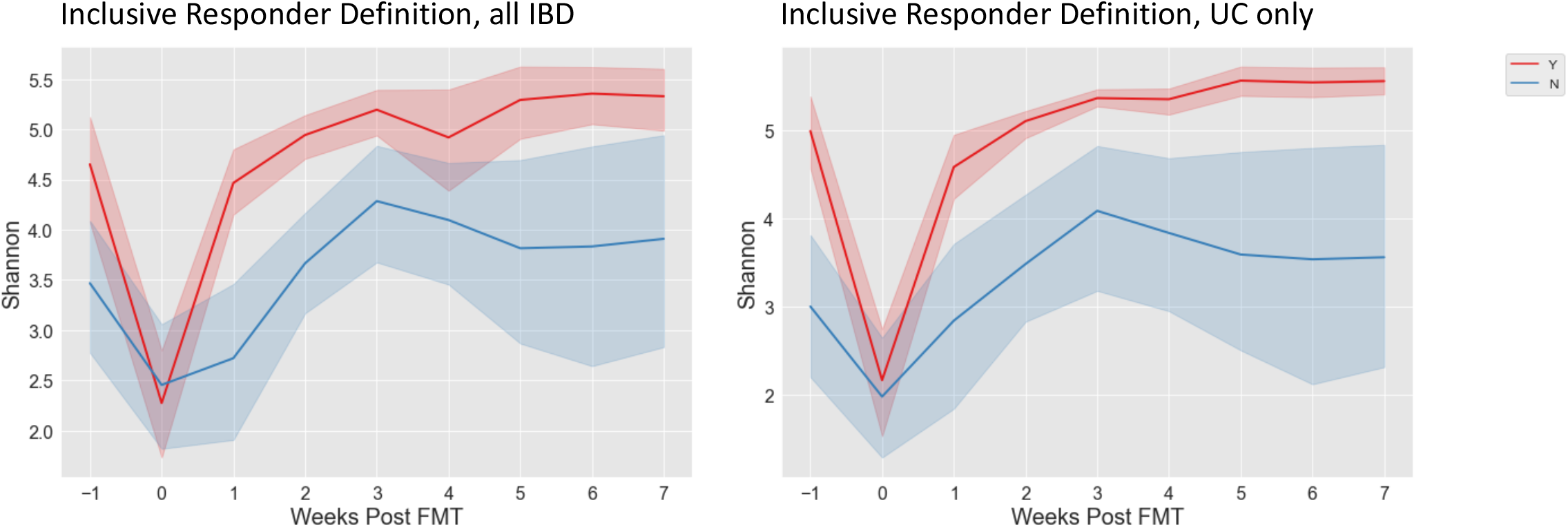
Shannon diversity of responders and non-responders using the more inclusive criteria for response for all IBD patients (left) and only UC patients (right).

**Supplemental Figure 2.**
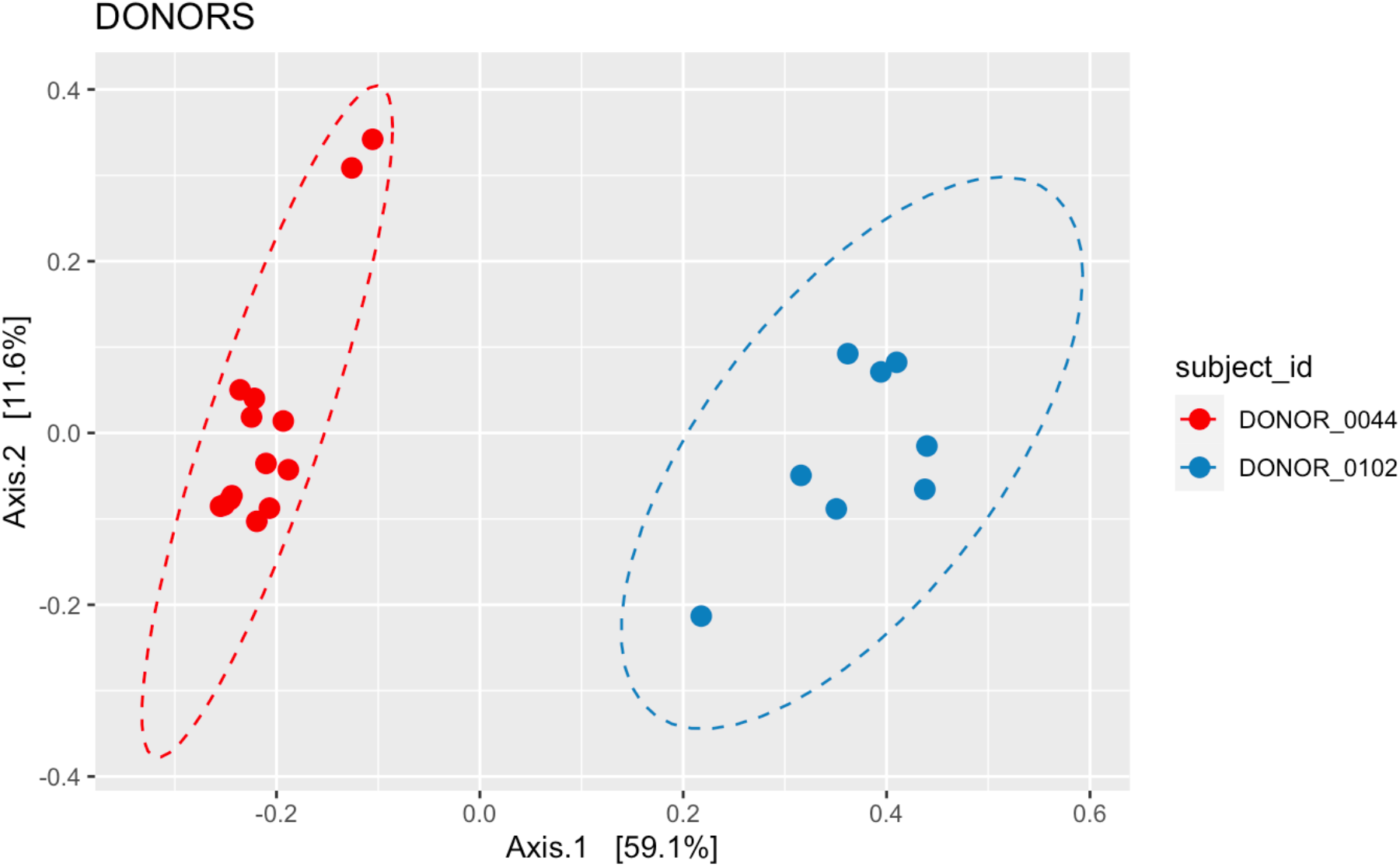
Principal Coordinate Analysis (Bray Curtis dissimilarity) for multiple stool samples from the two donors used in the study.

**Supplemental Figure 3.**
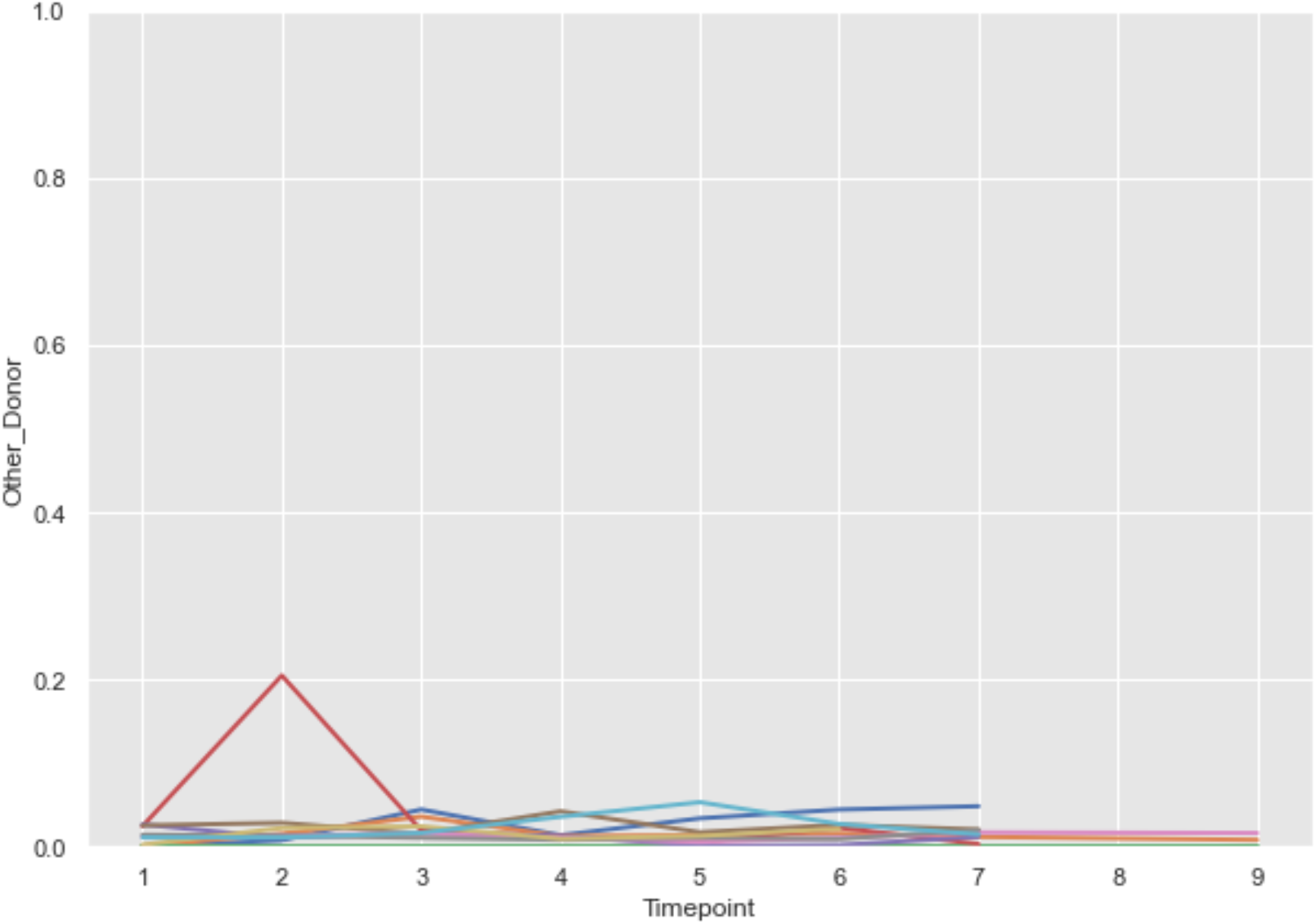
Percent of the post-FMT microbiome attributed to the “dummy donor” (the donor not used for FMT for any given subject was used as a potential source in SourceTracker). Each color represents a unique subject.

**Supplemental Figure 4.**
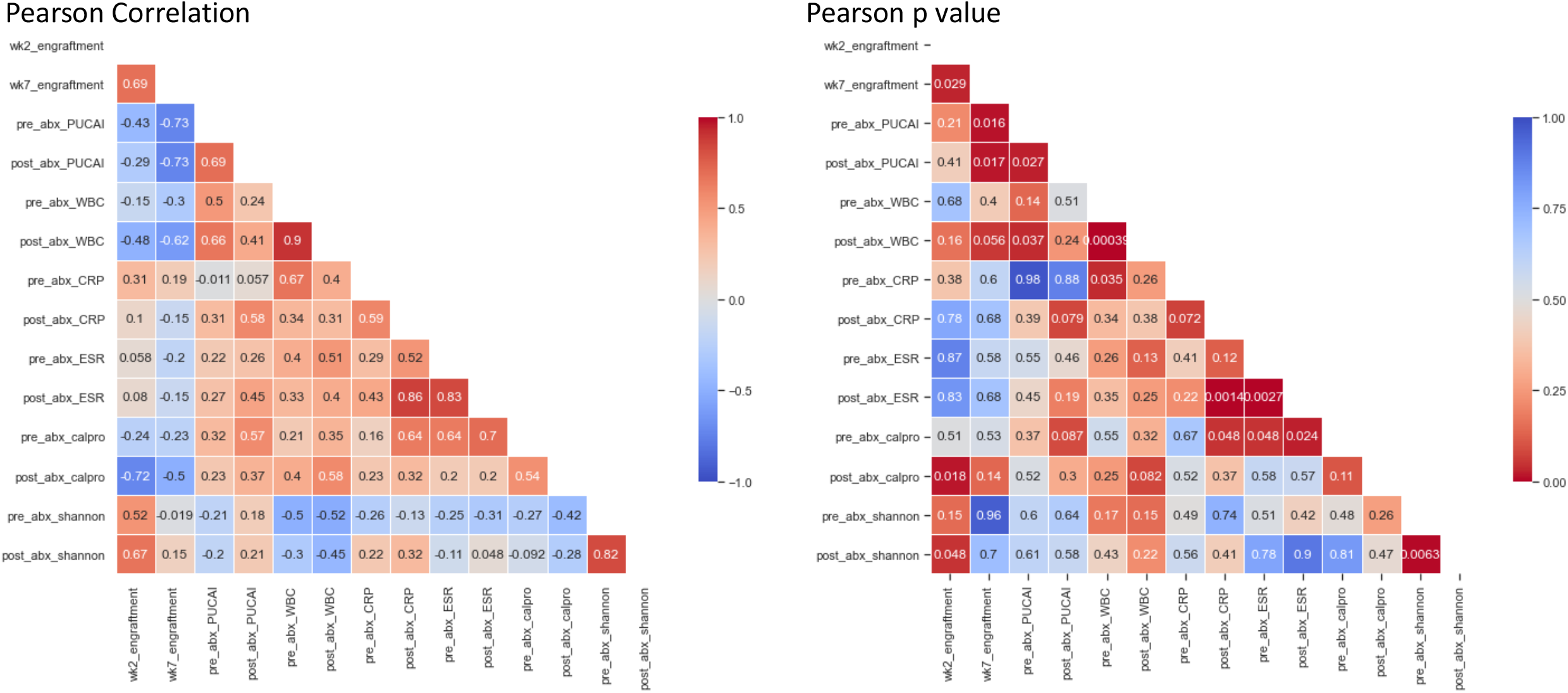
Correlation matrix of clinical, laboratory and microbiome factors related to engraftment. Pearson R (left) and P (right) values are shown.

**Supplemental Figure 5.**
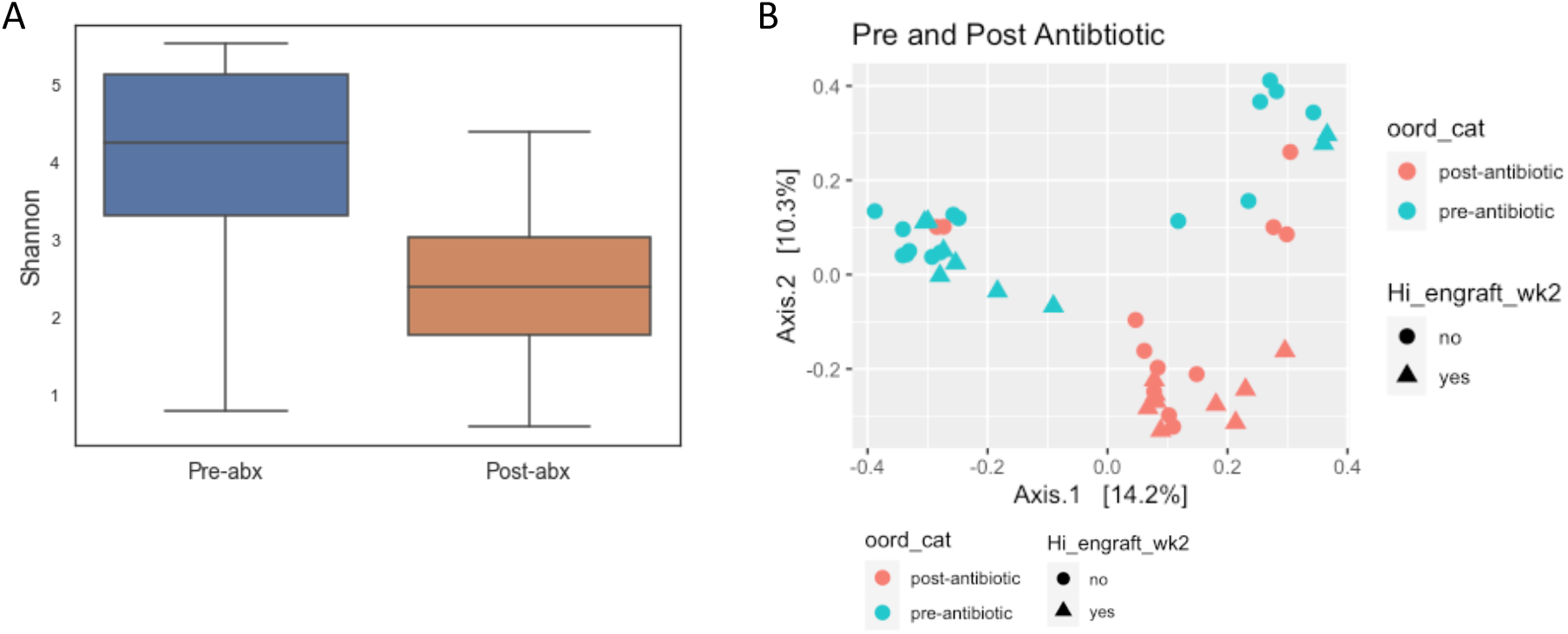
A. Shannon index pre- and post-antibiotics. B. Principal Coordinate Analysis of the pre and post antibiotic timepoints.

**Supplemental Figure 6.**
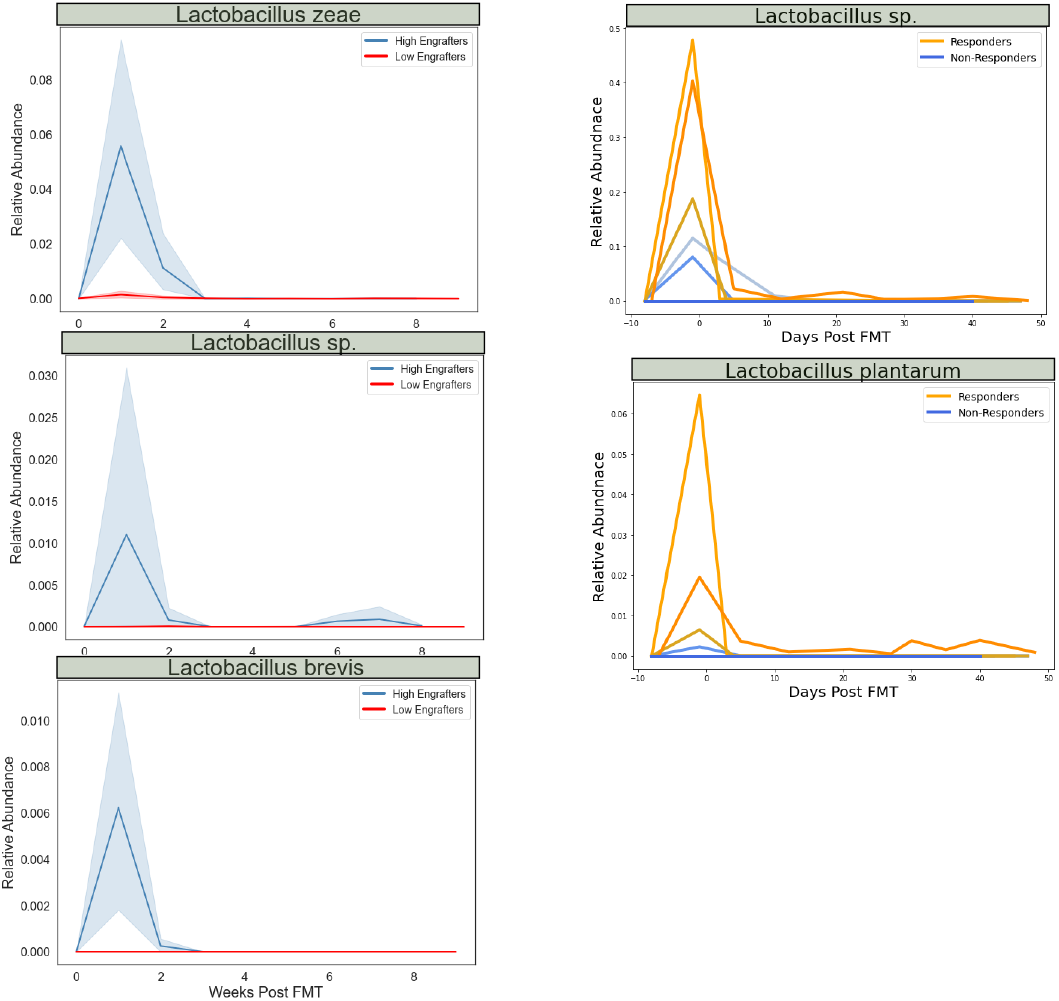
Lactobacillus relative abundance over time at the species level for both our study (left) and the reanalysis of Chu et al (right).

**Supplemental Table 1.** Clinical data table. Subject identifiers were randomly generated for the purposes of this manuscript and are not known to anyone outside the research team.

**Supplemental Table 2.** SourceTracker2 results. Subject identifiers were randomly generated for the purposes of this manuscript and are not known to anyone outside the research team

## Supplemental Text

### Adverse Event and Serious Adverse Event Reporting

There were 3 serious adverse events (SAE) that occurred. One CD patient had an episode of Grade 3 Colitis that was determined to be possibly related to the intervention. The subject was randomized to the FMT arm and was started on antibiotics. After completing the antibiotic regimen, the subject began experiencing a dramatic worsening of GI symptoms including bloody diarrhea, watery stools, and abdominal pain. The subject declined ED evaluation or hospitalization. Stool testing showed the subject was positive for a toxigenic *Clostridioides difficile* infection (CDI). The subject was started on vancomycin. Two days later, the subject’s rectal bleeding resolved, but continued to have watery stools and developed nausea. Despite an increase in vancomycin dosage, the patient had increasing abdominal pain, increased stools, nausea, vomiting, abdominal distention, fever and dehydration. At this time, the subject was evaluated in the ED and was found to have mild tachycardia and hypertension. Intravenous fluids and antibiotics were given and patient was admitted for 8-day inpatient stay until stable. This subject was removed from the study.

One UC patient (had a hypersensitivity reaction from the FMT induction enema. Minutes after the enema was given, the subject had dizziness, flushing, and chest tightness. Shortly after, the subject developed chills and Grade 3 sinus tachycardia. The patient remained tachycardic for 2 hours, at which point they were admitted to the emergency department (ED). In the ED, the subject had a high white blood cell count (WBC) and a low-grade fever. The subject was given antibiotics and admitted to the hospital. He was discharged 2 days later. This SAE was determined to be probably related to the FMT enema. This subject was removed from the study.

Another UC patient presented to the emergency department for worsening UC symptoms including increased abdominal pain, increased stool frequency, vomiting and dehydration. The subject was given intravenous fluids, total parenteral nutrition and an infusion of infliximab before being discharged. This occurred during the follow up period and was determined to be not related to the study intervention.

There were other Grade 1 and 2 adverse events including epiploic appendicitis, decreased white blood cell count, increased bilirubin, sore throat, upper respiratory infection, non-cardiac chest pain, gurgling bowel sounds, eye disorder, rash, eczema, flushing, anemia, increased alkaline phosphatase and an increased lymphocyte count; all were either not determined to be not related to the study intervention or were present at baseline. Most were resolved without sequelae.

---

**Table 1.**

|  | Crohn’s Disease Patients (n=4) | UC Patients (n=9) |
| --- | --- | --- |
| Sex | 3 Male | 9 Male |
| Race | 4 Caucasian | 9 Caucasian |
| Age at Diagnosis (Years) | 12 (Range: 8-16) | 19 (Range: 10-25) |
| Age at Enrollment (Years) | 20 (Range: 18-23) | 24 (Range: 14-29) |
| Disease Index |  |  |
| Mild | 4 | 3 |
| Mild/Moderate | 0 | 3 |
| Moderate | 0 | 3 |
| Concomitant Medications* |  |  |
| Amino-salicylates | 1 | 4 |
| Immunomodulators (6MP, MTX) | 0 | 2 |
| Anti-TNF Biologics | 3 | 4 |
| Other Biologics | 2 | 0 |
| Corticosteroids | 1 | 5 |
| Antibiotics | 2 | 2 |
| Baseline Fecal Calprotectin (µg/g)**<br>Normal: <50 | 1440 (Range: 1430-1450)<br>N=2 | 2190 (Range: 5-5730) |
| Baseline CRP (mg/L)**<br>Normal: 0-4.9 mg/L | 2.45 (Range: 0.3-5.2) | 1.9 (Range: 0.3-14.7) |
| Baseline ESR (mm/hr)**<br>Normal: 0-32 mm/hr | 14.5 (Range: 2-41) | 12 (Range: 2-79) |
| Prior IBD-Related Hospitalization |  |  |
| Yes | 2 | 6 |
| No | 0 | 1 |
| Unknown | 2 | 2 |
| Arm Assigned |  |  |
| Treatment | 2 | 6 |
| Placebo | 2 | 3 |

\*total > n because patients may be on multiple medications \*\*Median values shown

## Notes

### Clinical Trial

NCT02330653

### Author Declarations

The study was conducted in accordance with the protocol, applicable ICH Guidelines, Good Clinical Practice and the World Medical Association (WMA) Declaration of Helsinki and its amendments concerning medical research in humans. In accordance with guidelines and U.S. Code of Federal Regulations applicable to clinical studies, the protocol and informed consent/assent forms were reviewed and approved by the Boston Children's Hospital Institutional Review Board (IRB). The investigator informed the IRB and FDA of subsequent protocol amendments and reportable events as defined by IRB policy and FDA regulation. All patients and/or their guardians consented to participate in this study.

## References

1. Saha S, Mara K, Pardi DS, Khanna S. Long-term Safety of Fecal Microbiota Transplantation for Recurrent Clostridioides difficile Infection. Gastroenterology. 2021;160:1961-1969.e3.

2. Kelly CR, Yen EF, Grinspan AM, Kahn SA, Atreja A, Lewis JD, et al. Fecal Microbiota Transplantation Is Highly Effective in Real-World Practice: Initial Results From the FMT National Registry. Gastroenterology. 2021;160:183-192.e3.

3. Nicholson MR, Mitchell PD, Alexander E, Ballal S, Bartlett M, Becker P, et al. Efficacy of Fecal Microbiota Transplantation for Clostridium difficile Infection in Children. Clin Gastroenterol H. 2020;18:612-619.e1.

4. Cammarota G, Ianiro G. FMT for ulcerative colitis: closer to the turning point. Nat Rev Gastroentero. 2019;16:266–8.

5. Ishikawa D, Sasaki T, Osada T, Kuwahara-Arai K, Haga K, Shibuya T, et al. Changes in Intestinal Microbiota Following Combination Therapy with Fecal Microbial Transplantation and Antibiotics for Ulcerative Colitis. Inflamm Bowel Dis. 2017;23:116–25.

6. Crothers JW, Chu ND, Nguyen LTT, Phillips M, Collins C, Fortner K, et al. Daily, oral FMT for long-term maintenance therapy in ulcerative colitis: results of a single-center, prospective, randomized pilot study. Bmc Gastroenterol. 2021;21:281.

7. Chu ND, Crothers JW, Nguyen LTT, Kearney SM, Smith MB, Kassam Z, et al. Dynamic Colonization of Microbes and Their Functions after Fecal Microbiota Transplantation for Inflammatory Bowel Disease. Mbio. 2021;12:e00975–21.

8. Costello SP, Hughes PA, Waters O, Bryant RV, Vincent AD, Blatchford P, et al. Effect of Fecal Microbiota Transplantation on 8-Week Remission in Patients With Ulcerative Colitis: A Randomized Clinical Trial. Jama. 2019;321:156.

9. Rossen NG, Fuentes S, Spek MJ van der, Tijssen JG, Hartman JHA, Duflou A, et al. Findings From a Randomized Controlled Trial of Fecal Transplantation for Patients With Ulcerative Colitis. Gastroenterology. 2015;149:110-118.e4.

10. Vaughn BP, Vatanen T, Allegretti JR, Bai A, Xavier RJ, Korzenik J, et al. Increased Intestinal Microbial Diversity Following Fecal Microbiota Transplant for Active Crohn’s Disease. Inflamm Bowel Dis. 2016;22:2182–90.

11. Paramsothy S, Nielsen S, Kamm MA, Deshpande NP, Faith JJ, Clemente JC, et al. Specific Bacteria and Metabolites Associated With Response to Fecal Microbiota Transplantation in Patients With Ulcerative Colitis. Gastroenterology. 2019;156:1440-1454.e2.

12. Smillie CS, Sauk J, Gevers D, Friedman J, Sung J, Youngster I, et al. Strain Tracking Reveals the Determinants of Bacterial Engraftment in the Human Gut Following Fecal Microbiota Transplantation. Cell Host & Microbe. 2018;23:229-240.e5.

13. Narula N, Kassam Z, Yuan Y, Colombel J-F, Ponsioen C, Reinisch W, et al. Systematic Review and Meta-analysis. Inflamm Bowel Dis. 2017;23:1702–9.

14. Moayyedi P, Surette MG, Kim PT, Libertucci J, Wolfe M, Onischi C, et al. Fecal Microbiota Transplantation Induces Remission in Patients With Active Ulcerative Colitis in a Randomized Controlled Trial. Gastroenterology. 2015;149:102-109.e6.

15. Costello S, Waters O, Bryant R, Katsikeros R, Makanyanga J, Schoeman M, et al. OP036 Short duration, low intensity pooled faecal microbiota transplantation induces remission in patients with mild-moderately active ulcerative colitis: a randomised controlled trial. J Crohn’s Colitis. 2017;11:S23–S23.

16. Danne C, Rolhion N, Sokol H. Recipient factors in faecal microbiota transplantation: one stool does not fit all. Nat Rev Gastroentero. 2021;18:503–13.

17. Kellermayer R, Wu Q, Nagy-Szakal D, Queliza K, Ihekweazu FD, Bocchini CE, et al. Fecal Microbiota Transplantation Commonly Failed in Children With Co-Morbidities. J Pediatr Gastr Nutr. 2022;74:227–35.

18. Ianiro G, Punčochář M, Karcher N, Porcari S, Armanini F, Asnicar F, et al. Variability of strain engraftment and predictability of microbiome composition after fecal microbiota transplantation across different diseases. Nat Med. 2022;28:1913–23.

19. Smith BJ, Piceno Y, Zydek M, Zhang B, Syriani LA, Terdiman JP, et al. Strain-resolved analysis in a randomized trial of antibiotic pretreatment and maintenance dose delivery mode with fecal microbiota transplant for ulcerative colitis. Sci Rep-uk. 2022;12:5517.

20. Keshteli AH, Millan B, Madsen KL. Pretreatment with antibiotics may enhance the efficacy of fecal microbiota transplantation in ulcerative colitis: a meta-analysis. Mucosal Immunol. 2017;10:565–6.

21. Singh P, Alm EJ, Kelley JM, Cheng V, Smith M, Kassam Z, et al. Effect of antibiotic pretreatment on bacterial engraftment after Fecal Microbiota Transplant (FMT) in IBS-D. Gut Microbes. 2022;14:2020067.

22. Fischer M, Sipe B, Cheng Y-W, Phelps E, Rogers N, Sagi S, et al. Fecal microbiota transplant in severe and severe-complicated Clostridium difficile: A promising treatment approach. Gut Microbes. 2017;8:289–302.

23. Bolyen E, Rideout JR, Dillon MR, Bokulich NA, Abnet CC, Al-Ghalith GA, et al. Reproducible, interactive, scalable and extensible microbiome data science using QIIME 2. Nat Biotechnol. 2019;37:852–7.

24. Callahan BJ, McMurdie PJ, Rosen MJ, Han AW, Johnson AJA, Holmes SP. DADA2: High-resolution sample inference from Illumina amplicon data. Nat Methods. 2016;13:581–3.

25. McMurdie PJ, Holmes S. phyloseq: An R Package for Reproducible Interactive Analysis and Graphics of Microbiome Census Data. Plos One. 2013;8:e61217.

26. Pruesse E, Quast C, Knittel K, Fuchs BM, Ludwig W, Peplies J, et al. SILVA: a comprehensive online resource for quality checked and aligned ribosomal RNA sequence data compatible with ARB. Nucleic Acids Res. 2007;35:7188–96.

27. Knights D, Kuczynski J, Charlson ES, Zaneveld J, Mozer MC, Collman RG, et al. Bayesian community-wide culture-independent microbial source tracking. Nat Methods. 2011;8:761–3.

28. Bokulich NA, Dillon MR, Bolyen E, Kaehler BD, Huttley GA, Caporaso JG. q2-sample-classifier: machine-learning tools for microbiome classification and regression. J Open Res Softw. 2018;3:934.

29. Xie S, Zhao S, Jiang L, Lu L, Yang Q, Yu Q. Lactobacillus reuteri Stimulates Intestinal Epithelial Proliferation and Induces Differentiation into Goblet Cells in Young Chickens. J Agr Food Chem. 2019;67:13758–66.

